# Current and Lifetime Somatic Symptom Burden Among Transition-aged Autistic Young Adults

**DOI:** 10.1101/2021.10.02.21264461

**Authors:** Zachary J. Williams, Katherine O. Gotham

## Abstract

**Background and Objectives:** Somatic symptoms are the most common cause of outpatient medical visits in the general population, yet their presence and severity in individuals on the autism spectrum has rarely been studied. We sought to assess the prevalence, impact, and clinical correlates of fourteen commonly-reported somatic symptoms in a sample of transition-aged autistic young adults.

**Methods:** A sample of 290 independent and cognitively able autistic young adults (aged 18–26 years; mean [*SD*]: 23.10 [2.38] years) was recruited from the Simons Foundation SPARK participant pool. A modified version of the Patient Health Questionnaire–15 was used to assess somatic symptom prevalence/impact, along with measures of depression, anxiety, autistic traits, and quality of life.

**Results:** Somatic symptom burden was much higher in autistic young adults than previously reported in the general population. The most commonly reported current symptoms were fatigue (72.8%), sleep problems (69.0%), and menstrual problems (61.4% of females). Moderate or severe symptom levels were reported by 53.9% of females and 18.75% of males in our cohort, with the odds of females of endorsing any given symptom being 2–4 times greater than males. Both individual symptoms and total symptom burden were related to higher levels of depression, anxiety, and autistic traits, along with lower quality of life.

**Conclusion:** Despite little research on this topic previously, somatic symptoms are highly prevalent in autistic young adults, particularly women. Future research is needed to investigate links between somatic symptoms, medical and psychiatric morbidity, and health care utilization in the autistic population.

**What’s Known on This Subject:** Somatic symptoms are highly prevalent in the general population and account for a large proportion of health care costs. However, few studies have investigated the prevalence, impact, or correlates of these symptoms in individuals on the autism spectrum.

**What This Study Adds:** To our knowledge, this is the first study to specifically assess current and lifetime somatic symptom burden, symptom onset patterns, and the clinical correlates of multisystem symptom distress in transition-aged young adults on the autism spectrum.

## Introduction

As the diagnostic criteria for autism have grown broader, diagnoses of autism spectrum disorder (hereafter “autism”) have increased dramatically over the last three decades. Each year in the United States, approximately 50,000 individuals on the autism spectrum turn 18 years old, a number that is predicted to more than double over the next ten years^1^. Notably, transition-aged young adults on the autism spectrum generally utilize health care at higher rates relative to the general population and individuals with other neurodevelopmental disorders, with particularly high rates of primary care utilization^2^. In the general population, somatic symptoms represent the most common cause of outpatient medical visits, as well as the predominant reason why individuals with psychiatric conditions present to primary care settings^3^. However, with the notable exceptions of gastrointestinal symptoms^4^ and sleep disturbances^5,6^, somatic symptoms are infrequently studied in the autistic population^7–10^. Furthermore, no studies to date have investigated the prevalence of individual somatic symptoms or the natural history of these symptoms in autistic adults.

In this study, we used data from the Simons Powering Autism Research Knowledge (SPARK^11^) participant pool to assess self-reported current and lifetime somatic symptoms in a relatively large sample of transition-aged young adults on the autism spectrum. On the basis of previous studies in autistic individuals^7–10^ and the general population^12,13^, we hypothesized that young adults on the autism spectrum would report elevated levels of all somatic symptoms examined^7,8^, with higher somatic symptom burden associated with older age^10,12,13^, female sex^10,12,13^, higher levels of autistic traits^10^, elevated depression and anxiety symptoms^9,10,12,13^, and reduced quality of life^10,12,13^. In addition, we attempted to replicate results of a previous study wherein the relationship between autistic traits and somatic symptom burden was no longer significant after controlling for anxiety^10^. By describing the presence, severity, and clinical correlates of somatic symptoms in autistic individuals, the current study sought to further characterize these potentially major contributors to health care utilization and quality of life in this population.

## Methods

### Participants

Young adults from the United States with self-reported professional diagnoses of autism were recruited from the Simons Foundation-sponsored SPARK participant pool^11^ as part of a larger study on repetitive thinking and its clinical correlates in autistic adults (project number RM0030Gotham; see ^14^ for additional details). This larger study was advertised to all independent autistic adults between the ages of 18 and 45 in the SPARK participant pool (*n*=2887), 1014 of whom consented to complete the larger survey (35.1% response rate). As a part of this study, participants completed a series of self-report questionnaires via the SPARK platform on demographics, current and lifetime psychiatric diagnoses, autism severity, quality of life, co-occurring psychiatric symptoms, and a number of other clinical variables. Data for the current study were derived from the subset of individuals between 18 and 26 years of age who completed the portion of the study survey that included a measure of self-reported somatic symptom burden (*n*=290). All participants gave informed consent, and all study procedures were approved by the institutional review board at Vanderbilt University Medical Center.

### Measures

Somatic symptom burden was measured with a modified version of the Patient Health Questionnaire–15^15^, a well-validated somatization measure that is widely used in both medical and psychiatric settings. In the original PHQ-15, respondents rate the impact of 15 common symptoms (e.g., joint pain, dizziness, fatigue) over the past four weeks on a three-point Likert scale from 0 (*Not bothered at all*) to 2 (*Bothered a lot*). The current study used a modified version of the PHQ-15 (M-PHQ) that queried symptom burden at multiple time points, beginning with lifetime symptom presence (e.g., *Have you ever had problems with [symptom]?*), then asking participants to specify the developmental periods during which this symptom was problematic (i.e., childhood [0–12 years], adolescence [13–17 years], or adulthood [18+]), and finally to report the symptom’s impact over the previous three months (rated 0–2). This modified form also omitted the item assessing pain or problems during sexual intercourse, as part of a broader effort to reduce participant burden from our lengthy battery. As a measure of total somatic symptom burden across the remaining symptoms, we took the mean score of all non-missing items (i.e., up to 14 items for females and 13 for males), multiplied that score by 15, and rounded to the nearest integer value in order to approximate the 0–30 distribution of the original PHQ-15 questionnaire. Total scores on this measure can be qualitatively interpreted as minimal (0–4), mild (5–9), moderate (10–14), and severe (≥15)^15^.

Additional measures from the study survey battery were used in the current study to measure specific correlates of somatic symptom burden. Autistic traits were quantified using the self-reported Social Responsiveness Scale–Second Edition (SRS-2^16^) total T-score. Current symptoms of depression and generalized anxiety were measured using the Beck Depression Inventory–II (BDI-II) autism-specific latent trait score^14^ and the Generalized Anxiety Disorder–7 (GAD-7^15^) total score, respectively. Quality of life (QoL) was measured using the four-item World Health Organization Quality of Life scale previously validated in autistic adults (WHOQOL-4^17^). Item-level missing data were imputed using the mean of remaining items on each form.

### Statistical Analyses

Descriptive statistics were calculated for our sample, including the lifetime prevalence of each symptom, the prevalence of these symptoms during each developmental period, and the response frequencies for items assessing symptom impact. Corrected item-total polyserial correlations were also calculated for each symptom to assess its relationship with overall symptom burden. Current (3-month) and lifetime endorsement frequencies for each symptom were compared between male and female participants using odds ratios (ORs), and M-PHQ total scores were compared across sexes using Cohen’s *d*. Associations between M-PHQ scores and other outcomes were assessed using Pearson correlations, and polyserial correlations were used to assess relationships between individual symptoms and these same outcomes. Lastly, in order to determine the most important predictors of somatic symptom burden in this population, we regressed M-PHQ onto age, sex, SRS-2 score, and GAD-7 score, calculating partial *R*^2^ values to rank variable importance. A linear regression was used given the continuous nature of M-PHQ scores. Notably, BDI-II and GAD-7 scores were collinear, and thus only GAD-7 score was used in the regression due to content overlap between the BDI-II and PHQ-15 (i.e., sleep problems and fatigue appear on both questionnaires). Scale-level missing data was handled using 40-fold multiple imputation based on predictive mean matching. All statistical analyses were performed in R^18^.

## Results

Data were available for 290 young adults (61% female sex), with a mean (*SD*) age of 23.10 (2.38) years. Demographics and clinical characteristics of the participants, overall and divided according to biological sex, are presented in Table 1. The mean (*SD*) M-PHQ total score was 8.34 (6.02), with females reporting significantly higher symptom burden than males (*d*=0.906, 95% CI [0.657,1.15]). Moderate and severe levels of somatic symptoms were present in 23.4% and 16.9% of the total sample, respectively.

**Table 1.** Sociodemographic and Clinical Characteristics of the Sample.

|  | **Males**<br>**(*n* = 112)** | **Females**<br>**(*n* = 178)** | **Total**<br>**(*N* = 290)** | ***d* [95% CI]**<br>**(*F* > *M*)** |
| --- | --- | --- | --- | --- |
| Age (Years) | 22.65 (2.55) | 23.39 (2.23) | 23.10 (2.38) | 0.313 [0.074, 0.552] |
| Gender Identity |  |  |  |  |
| Cisgender | 103 (92.0%) | 146 (82.0%) | 249 (85.9%) |  |
| Transgender | 0 (0%) | 8 (4.5%) | 8 (2.8%) |  |
| Non-binary | 9 (8.0%) | 24 (13.5%) | 33 (11.4%) |  |
| Race/Ethnicity |  |  |  |  |
| Asian | 5 (4.5%) | 9 (5.1%) | 14 (4.8%) |  |
| Black/African American | 6 (5.4%) | 9 (5.1%) | 15 (5.2%) |  |
| Native American/Alaska Native | 1 (0.9%) | 12 (6.7%) | 13 (4.5%) |  |
| White | 82 (73.2%) | 145 (81.5%) | 227 (78.3%) |  |
| Other Race | 7 (6.2%) | 4 (2.2%) | 11 (3.8%) |  |
| Hispanic/Latino | 11 (9.8%) | 20 (11.2%) | 31 (10.7%) |  |
| Education |  |  |  |  |
| No High School Diploma | 5 (4.5%) | 7 (3.9%) | 12 (4.1%) |  |
| High School Diploma/GED | 43 (38.4%) | 39 (21.9%) | 82 (28.3%) |  |
| Vocational Certificate | 6 (5.4%) | 10 (5.6%) | 16 (5.5%) |  |
| Some College | 28 (25.0%) | 61 (34.3%) | 89 (30.7%) |  |
| Associate Degree | 7 (6.2%) | 15 (8.4%) | 22 (7.6%) |  |
| Bachelor's Degree | 18 (16.1%) | 35 (19.7%) | 53 (18.3%) |  |
| Graduate/Professional Degree | 5 (4.5%) | 11 (6.2%) | 16 (5.5%) |  |
| Age of Autism Diagnosis (Years) | 9.69 (6.32) | 13.71 (6.28) | 12.16 (6.58) | 0.637 [0.391, 0.883] |
| Ever had IEP for Autism | 79 (70.5%) | 86 (48.3%) | 165 (56.8%) |  |
| SRS-2 Total T-score | 66.68 (11.14) | 70.67 (10.70) | 69.14 (11.03) | 0.367 [0.125, 0.609] |
| BDI-II Autism-specific T-score | 47.32 (9.76) | 51.13 (9.19) | 49.66 (9.58) | 0.405 [0.164, 0.646] |
| GAD-7 Total Score | 6.51 (6.07) | 8.98 (5.79) | 8.02 (6.01) | 0.419 [0.177, 0.660] |
| WHOQOL-4 Total Score (1–5) | 3.43 (0.99) | 3.22 (0.85) | 3.30 (0.91) | -0.234 [-0.474, 0.005] |
| M-PHQ Total Score<sup>a</sup> | 5.28 (4.89) | 10.27 (5.87) | 8.34 (6.02) | 0.905 [0.657, 1.154] |
| Somatic Symptom Severity |  |  |  |  |
| Minimal (0–4) | 57 (50.9%) | 35 (19.7%) | 92 (31.7%) |  |
| Mild (5–9) | 34 (30.4%) | 47 (26.4%) | 81 (27.9%) |  |
| Moderate (10–14) | 16 (14.3%) | 52 (29.2%) | 68 (23.4%) |  |
| Severe (15+) | 5 (4.5%) | 44 (24.7%) | 49 (16.9%) |  |

*Note.* “Males” and “Females” refer to sex assigned at birth rather than gender. Values are presented as *M* (*SD*) for continuous variables and *N* (%) for categorical variables. *d* = Cohen’s *d* (standardized mean difference); IEP = Individualized Education Program; SRS-2 = Social Responsiveness Scale–Second Edition; BDI-II = Beck Depression Inventory–II; GAD-7 = Generalized Anxiety Disorder–7; WHOQOL-4 = Four-item World Health Organization Quality of Life Scale; M-PHQ = Modified Patient Health Questionnaire–15.
<sup>a</sup> Calculated as the 15 times the mean of all 14 items (13 for males), rounded to the nearest integer value.

Endorsement frequencies for each individual symptom are displayed in Table 2. The symptoms most highly endorsed in the past three months were fatigue (72.8%), sleep problems (69.0%), and menstrual problems (61.4% of females). Both current and lifetime symptom reports were higher in females for all symptoms (<u>Figure 2</u>), with confidence intervals excluding *OR*=1 for all but current syncope and lifetime dyspnea. The majority of participants reported that their symptoms were present since adolescence or adulthood, although abdominal pain, sleep problems, dyspnea, and constipation/diarrhea had an onset in childhood in over 40% of individuals reporting those symptoms. Item-total correlations were large for most symptoms, with dyspnea, chest pain, and abdominal pain showing the strongest relationships with overall symptom burden.

**Table 2.** Somatic Symptom Endorsement and Impact in 290 Transition-aged Autistic Young Adults.

| Symptom | Symptom Endorsement (%) | | | | | Three-month Symptom Impact (%) | | | $r_{it}$ |
| --- | --- | --- | --- | --- | --- | --- | --- | --- | --- |
|  | Current | Lifetime | Childhood<br>(0–12 yrs) | Adolescence<br>(13–17 yrs) | Adulthood<br>(18+ yrs) | Not<br>Bothered | Bothered<br>A Little | Bothered<br>A Lot |  |
| Fatigue | 72.8 | 80.3 | 19.0 | 58.6 | 76.9 | 27.2 | 33.4 | 39.3 | 0.572 |
| Sleep problems | 69.0 | 80.0 | 37.9 | 67.6 | 75.5 | 31.0 | 28.3 | 40.7 | 0.428 |
| Menstrual problems<sup>a</sup> | 61.4 | 82.4 | 5.2 | 44.5 | 44.8 | 23.4 | 21.4 | 15.9 | 0.151 |
| Back pain | 47.6 | 54.5 | 6.6 | 32.4 | 51.7 | 52.4 | 29.3 | 18.3 | 0.544 |
| Constipation/diarrhea | 46.6 | 56.9 | 27.9 | 46.2 | 50.7 | 53.4 | 26.9 | 19.7 | 0.567 |
| Nausea/gas/indigestion | 44.5 | 55.9 | 22.1 | 40.0 | 53.1 | 55.5 | 29.7 | 14.8 | 0.637 |
| Joint pain | 43.7 | 53.5 | 12.8 | 38.6 | 46.9 | 55.5 | 24.1 | 19.0 | 0.615 |
| Abdominal pain | 43.4 | 53.4 | 28.3 | 44.8 | 45.9 | 56.6 | 29.3 | 14.1 | 0.680 |
| Headache | 40.0 | 52.8 | 16.2 | 40.3 | 47.9 | 60.0 | 24.8 | 15.2 | 0.355 |
| Dizziness | 29.7 | 39.0 | 9.0 | 25.5 | 36.6 | 70.3 | 23.4 | 6.2 | 0.633 |
| Palpitations | 25.2 | 36.2 | 9.3 | 24.5 | 34.5 | 74.8 | 17.9 | 7.2 | 0.606 |
| Dyspnea | 24.2 | 33.9 | 15.9 | 26.2 | 29.3 | 75.5 | 17.9 | 6.2 | 0.718 |
| Chest pain | 17.6 | 30.3 | 6.6 | 19.3 | 24.5 | 82.4 | 15.5 | 2.1 | 0.687 |
| Syncope | 5.9 | 15.2 | 5.5 | 9.0 | 11.0 | 93.8 | 3.8 | 2.1 | 0.629 |

*Note.* Symptoms are displayed in order of current prevalence (past three months). Values indicate the percentage of the full sample endorsing each symptom or Likert scale response. $r_{it}$ = corrected item-total polyserial correlation for each symptom.
<sup>a</sup> Based on results from female participants only.

Correlations between PHQ symptoms, total scores, and covariates of interest are displayed in Table 3. As expected, total symptom burden was strongly related to depression (*r*=0.522 [0.432,0.601]) and anxiety (*r*=0.512 [0.421,0.593]) symptoms, displaying slightly smaller correlations with autistic trait levels (*r*=0.432 [0.333,0.522]) and QoL (*r*=-0.323 [-0.423, -0.216]). A small but significant correlation was also present between M-PHQ total score and age, with older individuals reporting slightly more symptoms (*r*=0.145 [0.030,0.256]). Correlations at the individual symptom level were attenuated but still statistically significant for all symptoms except menstrual problems (Table 3).

**Table 3.** Correlations of Individual Somatic Symptoms with External Variables.

| Symptom | BDI-II | GAD-7 | SRS-2 | WHOQOL-4 | Age |
| --- | --- | --- | --- | --- | --- |
| Fatigue | 0.659<br>[0.589, 0.720] | 0.479<br>[0.385, 0.563] | 0.315<br>[0.208, 0.415] | -0.412<br>[-0.503, -0.311] | 0.107<br>[-0.008, 0.220] |
| Sleep problems | 0.487<br>[0.394, 0.570] | 0.418<br>[0.318, 0.509] | 0.411<br>[0.311, 0.502] | -0.352<br>[-0.449, -0.247] | 0.164<br>[0.050, 0.274] |
| Menstrual problems | 0.076<br>[-0.039, 0.190] | 0.165<br>[0.050, 0.275] | 0.092<br>[-0.023, 0.205] | 0.068<br>[-0.047, 0.182] | 0.040<br>[-0.076, 0.154] |
| Back pain | 0.454<br>[0.358, 0.541] | 0.390<br>[0.288, 0.483] | 0.299<br>[0.191, 0.401] | -0.248<br>[-0.353, -0.136] | 0.116<br>[0.000, 0.228] |
| Constipation/diarrhea | 0.293<br>[0.184, 0.395] | 0.343<br>[0.237, 0.441] | 0.227<br>[0.114, 0.333] | -0.123<br>[-0.235, -0.008] | 0.116<br>[0.000, 0.228] |
| Nausea/gas/indigestion | 0.237<br>[0.125, 0.343] | 0.291<br>[0.182, 0.393] | 0.232<br>[0.120, 0.338] | -0.124<br>[-0.236, -0.009] | 0.201<br>[0.088, 0.309] |
| Joint pain | 0.328<br>[0.221, 0.427] | 0.383<br>[0.281, 0.478] | 0.350<br>[0.245, 0.447] | -0.278<br>[-0.381, -0.168] | 0.026<br>[-0.090, 0.141] |
| Abdominal pain | 0.301<br>[0.192, 0.402] | 0.344<br>[0.238, 0.442] | 0.313<br>[0.205, 0.413] | -0.149<br>[-0.260, -0.035] | 0.124<br>[0.009, 0.236] |
| Headache | 0.291<br>[0.182, 0.393] | 0.357<br>[0.252, 0.453] | 0.388<br>[0.286, 0.482] | -0.180<br>[-0.289, -0.066] | 0.105<br>[-0.011, 0.217] |
| Dizziness | 0.440<br>[0.342, 0.528] | 0.370<br>[0.266, 0.465] | 0.382<br>[0.279, 0.476] | -0.319<br>[-0.419, -0.212] | 0.068<br>[-0.047, 0.182] |
| Palpitations | 0.405<br>[0.304, 0.497] | 0.376<br>[0.273, 0.471] | 0.316<br>[0.208, 0.416] | -0.275<br>[-0.378, -0.165] | 0.153<br>[0.038, 0.264] |
| Dyspnea | 0.265<br>[0.155, 0.369] | 0.257<br>[0.146, 0.361] | 0.246<br>[0.135, 0.351] | -0.156<br>[-0.266, -0.042] | -0.006<br>[-0.121, 0.109] |
| Chest pain | 0.233<br>[0.121, 0.339] | 0.238<br>[0.127, 0.344] | 0.194<br>[0.081, 0.303] | -0.203<br>[-0.311, -0.090] | -0.036<br>[-0.150, 0.080] |
| Syncope | 0.520<br>[0.431, 0.599] | 0.453<br>[0.356, 0.540] | 0.443<br>[0.346, 0.531] | -0.356<br>[-0.453, -0.251] | -0.041<br>[-0.155, 0.075] |

*Note.* Polyserial correlations are presented along with 95% confidence intervals. BDI-II = Beck Depression Inventory-II; GAD-7 = Generalized Anxiety Disorder-7; SRS-2 = Social Responsiveness Scale-Second Edition; WHOQOL-4 = Four-item World Health Organization Quality of Life Scale

Regression analyses indicated that the combination of age, sex, SRS-2 scores, and GAD-7 scores explained a sizable portion of variance in overall somatic symptom burden (*R*^2^=0.390). Notably, after controlling for sex, anxiety, and autistic traits, age was no longer associated with symptom burden (β_Std_ =0.008 [-0.087,0.104], *R*^2^_p_ <0.001). However, the remaining three predictors all explained a significant portion of unique variance in M-PHQ scores, with female sex (β=0.608. [0.413,0.804], *R*^2^_p_ =0.081) and GAD-7 score (β_Std_ =0.365 [0.253,0.476], *R*^2^_p_ =0.090) explaining a larger proportion of the variance than SRS-2 score (β_Std_ =0.184 [0.071,0.296], *R*^2^_p_ =0.022).

**Figure 1.**
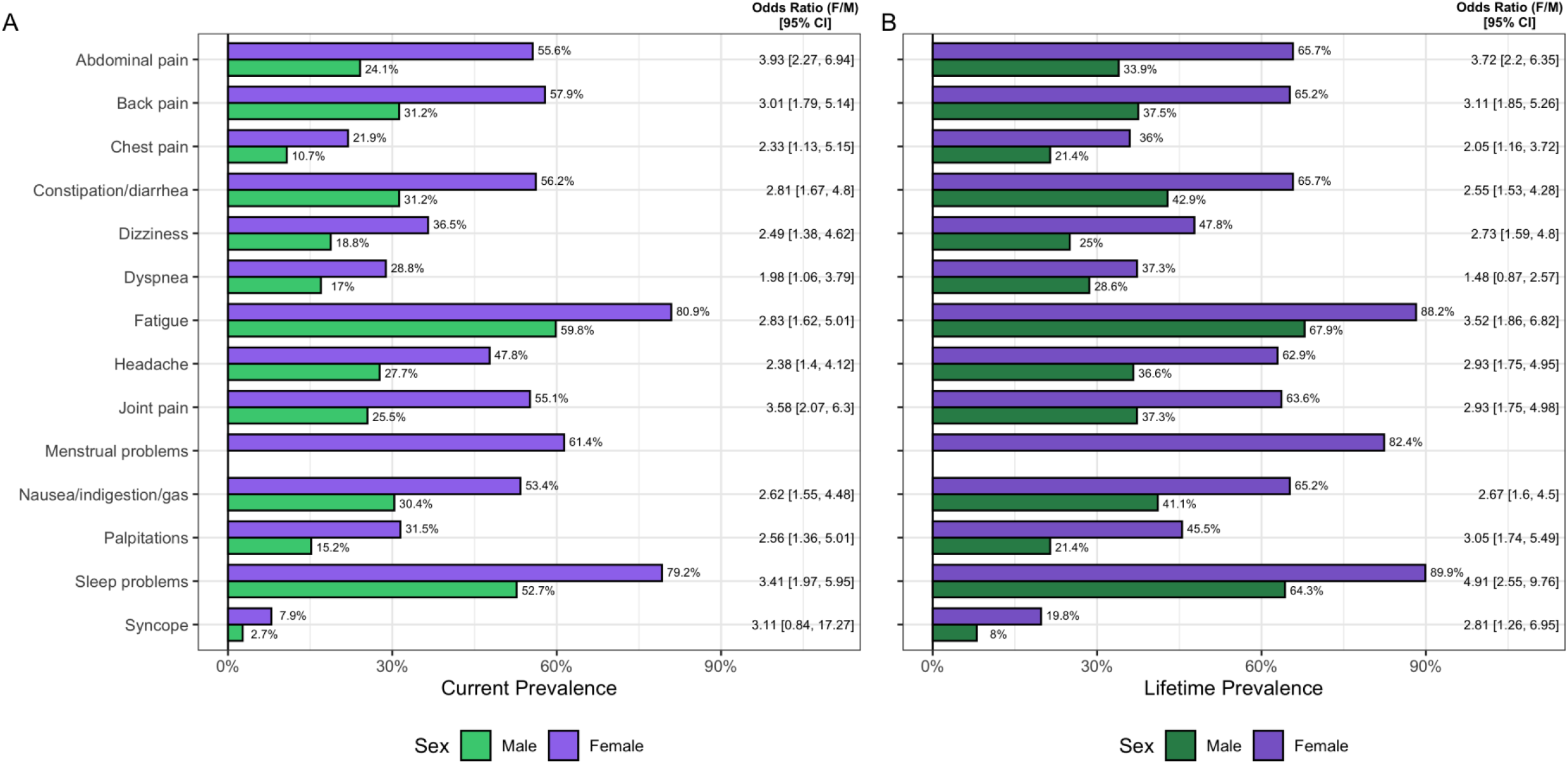
Sex differences in **(A)** current endorsement (“a little” or “a lot” bothered in past three months) and **(B)** lifetime endorsement (“ever” bothered by symptom) of fourteen somatic symptoms by autistic young adults. Odds ratios (Female/Male) are presented for each symptom along with 95% maximum likelihood confidence intervals.

## Discussion

In this study, we examined the current and lifetime prevalence of fourteen common somatic symptoms in a cohort of transition-aged autistic young adults, finding that these individuals endorsed all somatic symptoms at much higher rates than young adults in previous general population studies using the PHQ-15^13,19^. The majority of these somatic symptoms were reported to have an onset in adolescence or adulthood, although gastrointestinal complaints, sleep problems, and dyspnea appeared in childhood for many individuals reporting those symptoms. Covariation between these symptoms was very strong, with 11 of 14 symptoms exhibiting corrected item-total polyserial correlations greater than 0.5. Moreover, when considering total somatic symptom burden, 40.3% of autistic young adults (53.9% of females and 18.8% of males) reported total symptoms in the “moderate” or “severe” range. Overall somatic symptom burden was strongly associated with female sex, more severe symptoms of anxiety and depression, higher levels of autistic traits, and lower overall QoL. Somatic symptom burden was slightly related to older age, but this relationship disappeared in regression analyses, indicating it was likely due to the slight age difference between males and females in our sample. Additionally, the positive relationship between autistic traits and somatic symptom burden remained after controlling for age, sex, and generalized anxiety, in contrast to prior findings in an adult autistic sample^10^. However, it is notable that the prior study also controlled for self-reported sensory sensitivity (Sensory Perception Quotient scores), which were not measured in the current study. Additionally, measures of autistic traits, somatic symptom burden, and anxiety differed between the two studies, potentially leading to diverging results. Additional studies are therefore necessary to investigate this discrepancy and determine whether the demonstrated relationship between autistic traits and somatic symptom burden can indeed be explained entirely by co-occurring psychopathology.

As with prior general population^12,13,19^ and autistic adult^10^ samples, we found large sex differences in somatic symptom reporting, with the odds of reporting a given symptom in the past three months being 2–4 times higher in female compared to male participants. These results are consistent with the higher rates of co-occurring physical health conditions seen in autistic females compared to males^20^. The sex difference in total symptom burden was also very large (*d*>0.9), with a magnitude greater than double that seen in the general population^12^. This sex difference also remained large in the multiple regression analysis, which controlled for observed sex differences in age, autistic traits, and anxiety symptomatology. These findings stand in contrast to the lack of sex differences in somatic complaints reported in a primarily prepubescent sample of children on the autism spectrum^8^. As the majority of somatic symptoms in the current sample were reported to have arisen in adolescence or adulthood, it is therefore possible that sex differences in somatic symptom burden may only emerge after puberty, as has been observed in the general population^21^. Longitudinal studies in autistic youth are necessary to accurately characterize the onset, persistence, and developmental trajectory of multiple somatic symptoms in this population, additionally determining whether sex differences in these symptoms are limited to certain developmental periods.

Limitations of our study include a female-predominant and therefore unrepresentative sample of autistic adults, a relatively low response rate to the study survey, no neurotypical control participants who completed the same M-PHQ, a lack of determination regarding which symptoms were attributable to diagnosed somatic conditions, and an inability to relate symptoms to health anxiety or health care utilization. In particular, given the large sex differences in somatic symptom burden, the 40% rate of clinically significant somatization in our sample is likely an overestimate of the true population prevalence, with sex-weighted prevalence estimates of 25.8% and 27.5% based on our data using 4:1 and 3:1 male to female ratios, respectively^22^. An additional limitation concerns the interpretation of scores from the M-PHQ, which included a longer recall period compared to the original PHQ-15 and may thus overestimate current symptom endorsement slightly. Moreover, as normative values for the M-PHQ were not investigated in a general population sample, it is possible that the prevalence of some somatic symptoms in this sample does not substantially exceed that reported by general population controls. Somatic symptom questionnaires such as the PHQ-15 also do not assess the etiology of the experienced symptoms, and thus we were unable to determine the degree to which these symptoms are medically explainable. As autistic individuals have increased risk of medical conditions such as inflammatory bowel disease^4^ and sleep-disordered breathing^5,6^, it is important that practitioners do not simply forego appropriate medical screenings because symptoms in this population are presumed to be functional in origin. Future work is therefore necessary to investigate the degree to which organic pathology contributes to the burden of somatic symptoms in autistic individuals. Lastly, the current study was limited to young adults on the autism spectrum; much less work has been conducted with middle-aged and older autistic adults^7^, and it is currently unclear whether the prevalence, burden, and correlates of somatic symptoms in the autistic population change over the adult lifespan.

## Conclusions

Somatic symptoms are among the most common reasons for health care utilization in the general population, yet these symptoms have been largely overlooked in the study of physical and mental health in autistic individuals^10^. In a large sample of transition-aged autistic young adults, the current study reported a substantially elevated prevalence of somatic symptoms across multiple organ systems, additionally demonstrating relationships between total symptom burden and anxiety, depression, autistic trait levels, and quality of life. Our findings should be interpreted with caution, as they are based on a predominantly female, later-diagnosed sample of cognitively able adults on the autism spectrum whose diagnoses were not confirmed clinically. Thus, these findings should be replicated in more rigorously-phenotyped samples with additional characterization of co-occurring physical and psychiatric symptoms and diagnoses. Although the effects of somatic symptoms on health care utilization and overall morbidity in the autistic population are still undetermined, our findings indicate that consideration of somatic symptoms is warranted when investigating the health status of individuals on the autism spectrum.

## Data Availability

Approved researchers can obtain the SPARK population dataset described in this study (project number RM0030Gotham) by applying at https://base.sfari.org. 

## Abbreviations

BDI-II: Beck Depression Inventory–II
GAD-7: Generalized Anxiety Disorder–7
M-PHQ: Modified version of the Patient Health Questionnaire–15
PHQ-15: Patient Health Questionnaire–15
QoL: Quality of Life
SPARK: Simons Powering Autism Research Knowledge
SRS-2: Social Responsiveness Scale–Second Edition
WHOQOL-4: World Health Organization Quality of Life–4 item scale

## Acknowledgements

The authors are grateful to all of the individuals and families enrolled in SPARK, the SPARK clinical sites and SPARK staff. They appreciate obtaining access to demographic and phenotypic data on SFARI Base. Approved researchers can obtain the SPARK population dataset described in this study (project number RM0030Gotham) by applying at https://base.sfari.org.

